# Common Practices for Sociodemographic Data Reporting in Digital Mental Health Intervention Research: A Scoping Review

**DOI:** 10.1101/2023.07.24.23293020

**Authors:** Andrew Kirvin-Quamme, Jennifer Kissinger, Laurel Quinlan, Robert Montgomery, Mariya Chernenok, Maddison C. Pirner, Sarah Pajarito, Stephanie Rapoport, Paul Wicks, Alison Darcy, Carolyn J. Greene, Athena Robinson

**Affiliations:** Woebot Health, San Francisco, CA, USA; Wicks Digital Health, Ltd. Lichfield, UK; Translational Research Institute (TRI), University of Arkansas for Medical Sciences (UAMS)

**Keywords:** Scoping review, digital mental health interventions, app-based, RCTs, clinical trials, sociodemographic, demographic, treatment disparities, marginalized groups, self-identity

## Abstract

**Background:** The ability for digital mental health interventions (DMHI) to reduce mental health disparities relies on recruitment of research participants with diverse sociodemographic and self-identity characteristics. Despite its importance, sociodemographic reporting in research is often limited, and the state of reporting practices in DMHI research in particular has not been comprehensively reviewed.

**Objectives:** To characterize the state of sociodemographic data reported in randomized controlled trials (RCTs) of app-based DMHIs published globally from 2007 to 2022.

**Methods:** A scoping review of RCTs of app-based DMHIs examined reporting frequency for 16 sociodemographic domains (i.e., Gender) and common category options within each domain (i.e., woman). The search queried five electronic databases. 5079 records were screened and 299 articles were included.

**Results:** On average, studies reported 4.64 (SD = 1.79; range 0 - 9) of 16 sociodemographic domains. The most common were Age (97%) and Education (67%). The least common were Housing Situation (6%), Residency/Location (5%), Veteran Status (4%), Number of Children (3%), Sexual Orientation (2%), Disability Status (2%), and Food Security (<1%). Gender or Sex was reported in 98% of studies: Gender only (51%), Sex only (28%), both (<1%), Gender/Sex reported but unspecified (18%). Race or Ethnicity was reported in 48% of studies: Race only (14%), Ethnicity only (14%), both (10%), Race/Ethnicity reported but unspecified (10%).

**Conclusions:** This review describes widespread underreporting of sociodemographic information in RCTs of app-based DMHIs published from 2007 to 2022. Reporting was often incomplete (i.e., % female only), unclear (i.e., conflation of Gender/Sex), and limited (i.e., only options representing majority groups were reported). Trends suggest reporting somewhat improved in recent years. Diverse participant populations must be welcomed and described in DMHI research to broaden learnings and generalizability of results; a prerequisite of DMHI’s potential to reduce disparities in mental healthcare.

**STRENGTHS AND LIMITATIONS OF THIS STUDY:**

- This study is the first of its kind to assess global sociodemographic reporting practices in RCTs of treatment outcomes research for app-based DMHIs
- This review was both large and comprehensive, screening over 5000 articles leading to the inclusion of nearly 300 articles spanning the entire lifespan of app-based DMHIs and extracting data for a wide array of 16 sociodemographic domains
- Article inclusion criteria allowed for a broad range of DMHI studies, including populations with both clinical and sub-clinical conditions
- It was not feasible to report on the sociodemographic composition of each study sample due to the large number of studies included in the review and the breadth of domains evaluated
- This review is descriptive, and did not formally assess statistical differences in reporting practices between different sub-groups or time periods, or provide any assessment of barriers, facilitators, or solutions to the issues identified

## BACKGROUND

Mental health, like physical health, is ubiquitous, regardless of identity. However, marginalized individuals, including those who have been and/or are actively discriminated against and experience social, political, and economic exclusion due to factors such as age, race, ethnicity, gender, sexual orientation, religion, disability, and socioeconomic status, are frequently impacted by mental health inequities (1–3). Such inequities may include experiencing higher acute and/or lifetime prevalence rates of depression, anxiety, post-traumatic stress disorder, substance use, and suicide (4–14), as well as being more likely to be un- or under-diagnosed and less likely to receive treatment for their presenting mental health concern (9,15,16). It is widely acknowledged that clinical psychology research has played a vital role in not only facilitating our current understanding of mental health conditions, their prevalence and course, but also in identifying and refining evidence based mental health treatment protocols that can meaningfully improve patient symptoms and quality of life (17). Unfortunately, such research has a longstanding history of limited sociodemographic diversity among its samples (18–20). The impact of limited diversity in sample composition has substantial downstream effects. Homogenous research samples – those lacking in diverse ethnicities, races, ages, genders, sexual orientations, and socioeconomic backgrounds, or lacking data regarding these characteristics – limit opportunities to develop and test interventions of interest among such populations, thereby reducing both clinical research learnings and generalizability of findings. Over time, the chronic lack of diversity and inclusion in such research may further alienate underrepresented groups and inhibit them from volunteering their participation in such studies, increase wariness of the medical research system, and diminish opportunity to receive care via clinical research or healthcare ecosystems (21,22). Conversely, the purposeful and directed inclusion of participants from various backgrounds provides opportunities to identify similarities or differences in treatment response, utilization, satisfaction, side effects, and more. This in turn, illuminates treatment adaptation opportunities when indicated (23) and ultimately encourages the generation of a scientific literature that is fundamentally more representative of all peoples. Despite the knowledge of the significant impact of sociodemographic domains on mental health and associated outcomes, and the importance of recruiting diverse samples for addressing the numerous disparities in these outcomes, the actual reporting of sociodemographic information in mental health research is often limited (24–26). One possible reason for limited sociodemographic reporting may be due to researchers’ efforts to reduce participant’s assessment burden by limiting the number of questionnaire items pertaining to sociodemographic information (27). Another possible reason is a lack of standardized approaches to collecting sociodemographic information in mental health research. While several guidelines have been published, some are outdated and discordant to current cultural norms, few offer best practices for how to collect this information, and none are comprehensive enough to include all relevant sociodemographic factors (28–33). A third may be related to restrictions imposed by scientific journals on word count, table size, or other formatting requirements, which may cause researchers to reduce or selectively report sociodemographic information that was actually collected.

Digital mental health interventions (DMHIs) present an opportunity to improve both structural (e.g., cost, lack of or insufficient insurance coverage, provider shortages, service inaccessibility, and inadequate services) and attitudinal (e.g., perceived stigma) barriers to care (2,34–36). Indeed, DMHIs have enormous potential to improve service accessibility in marginalized communities; however, individuals from these populations may be consistently under-represented in DMHI research (37–39), unfortunately emulating the larger clinical research field. The ability for DMHIs to be optimally poised to minimize barriers contributing to mental health disparities is attenuated if DMHI research continues to analyze homogenous sociodemographic and self-identity samples. However, efforts to diversify samples rely on comprehensive collection and reporting of sociodemographic information. To date, the state of sociodemographic reporting practices in DMHI research has not been comprehensively reviewed. Therefore, the present study aims to characterize sociodemographic and self-identity data reporting practices in DMHI outcomes research. This review may provide a foundational component for the development of comprehensive sociodemographic and self-identity data reporting guidelines for future research.

### Primary Objectives

To investigate the common practices for the reporting of sociodemographic information in randomized controlled trials (RCTs) of app-based DMHIs (henceforth “DMHIs”) published globally from 2007-2022. The authors sought to address this overarching aim via two specific research questions:

1. What are the common sociodemographic domains reported in RCTs of DMHIs and how frequently is each sociodemographic domain reported?
2. What are commonly reported category options within each sociodemographic domain?

### Secondary Objectives

Sociodemographic reporting trends may change over time. The National Institute of Health’s (NIH) guidelines regarding sociodemographic domains were published in 2016 (40) and updated in 2017 (41) and 2019 (31), and other studies have found a general increase in reporting of sociodemographic variables over the past four decades (24). Additionally, sociodemographic reporting practices are not internationally standardized, and different countries may develop unique practices suited to their context. As such, secondary objectives were to describe reporting practices across time, and to separate United States (US) and non-US studies in order to isolate reporting practices in the US specifically. These included the specific research questions:

1. Does the reporting frequency of sociodemographic domains appear to change over time (i.e., from 2007 to 2022)
2. Does the reporting of sociodemographic domains appear to differ between US and non-US countries?

## METHODS

### Protocol

The study protocol was developed using the Joanna Briggs Institute (JBI) methodology for scoping reviews (42) and was pre-registered on the Open Science Framework (https://osf.io/rs3c4). There were several deviations from the protocol. First, we did not conduct a search of all references within each included source of evidence as originally planned, as the original search returned more included studies than anticipated. Additional screening would have exceeded the research team’s bandwidth. Second, the protocol originally planned to assess the extent to which studies in the review allowed for self expression; for example, by providing the ability to select multiple options for one sociodemographic domain, the ability to write in a response, as well as the phrasing of write-in options (e.g., “other”). Unfortunately, studies did not provide consistent or clear information on these questions, and therefore we were unable to code for their presence. Third, Google Sheets, rather than JBI SUMARI, was used to manage coding, due to technical difficulties with JBI SUMARI. Fourth, exclusion reasons were not coded at the title and abstract review stage because this is not a Preferred Reporting Items for Systematic Reviews and Meta-Analyses (PRISMA) requirement. Fifth, after the publication of the protocol, the domain “Residency/Location” was added in order to capture whether a sample was described as rural, urban, etc.

### Eligibility Criteria

Manuscripts that reported on RCTs of app-based DMHIs were included. As RCTs are considered the “gold standard” of clinical research (43), data collection and reporting practices in RCTs are expected to be the most rigorous of any clinical research design. Therefore, all other study designs were excluded in alignment with this review’s objective to characterize sociodemographic data reporting practices for a relatively homogenous set of studies. The DMHIs must have been primarily designed 1) to help patients with mental health disorders (e.g. depression) or well-being concerns (e.g. stress) and 2) for use on a personal mobile device delivered through a downloadable smartphone app (e.g. in App Store and Google Play) that could be used outside of a clinic setting. The articles must have been available in English and published in a peer-reviewed journal (i.e., not a conference abstract or dissertation). No restrictions were placed on the country of study origin or RCT control group type. Articles published from January 1, 2007 onward were included in the review, reflecting the launch of the first major mobile app store in 2007. DMHIs with an adjunct or combined therapeutic intervention were included as long as the primary intervention remained the app-based component. We excluded studies not primarily focused on app-based DMHIs, including virtual reality, computer-delivered, or video/phone-delivered psychotherapies and interventions delivered primarily through messaging-based apps (i.e., WeChat and WhatsApp). To avoid double-counting studies which produced multiple papers, only primary studies were included in this review.

### Information Sources

The search queried five databases: MEDLINE, CINAHL, PsycINFO, Google Scholar, and PsyArXiv. A three-step search strategy was utilized to develop the keyword search string in this scoping review per JBI guidelines (42). First, a search of MEDLINE and CINAHL was conducted to identify articles on this topic. The keyword string for our initial search was developed by reviewing search strings used in systematic literature reviews cited within a recent meta-analysis of mobile DMHIs (44). Second, a comprehensive and non-repetitive list of relevant keywords used inclusively in these studies (44) was used to develop the keyword search string for the preliminary search. Third, a preliminary search was conducted to identify additional keywords not included from the previous steps. Following recommendations by JBI (42) to ensure a comprehensive keyword string, on August 3, 2022 the preliminary search was conducted in MEDLINE and CINAHL for the first three results meeting the inclusion criteria. Titles and abstracts from these first three citations were reviewed for relevant keywords. Modifications to the original search string from step one and two incorporated terms found within the first three citations that were not included in the original search string. With the addition of new terms (psychological, psychology, behavioral, mindfulness, oppositional defiant disorder, fatigue, and self-compassion) the keyword list for the full search string was considered final (see Supplemental 2).

### Search Strategy

The search was conducted on August 9, 2022. Following recommendations from Haddaway and colleagues, Google Scholar results were limited to the first 200 citations (45). Citations from the full search were uploaded into individual Zotero folders based on the search database, with duplicates removed during the import process to Zotero (Zotero 6.0.4, 2022) and export process (EBSCO). A flow diagram of the search results is reported in Figure 1.

**Figure 1.**
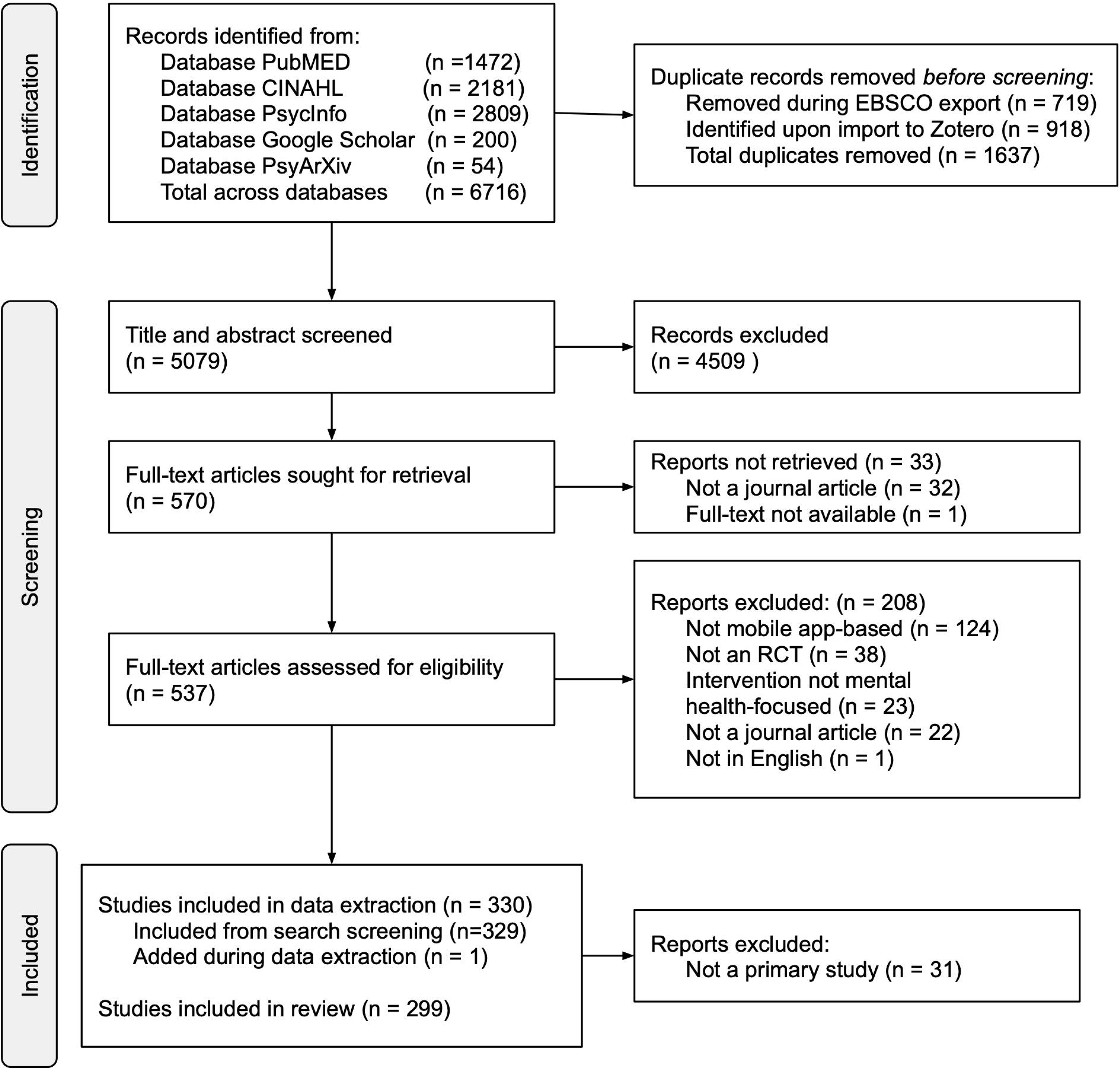
PRISMA flow diagram of study inclusion.

### Study Selection Process

Prior to commencing screening, two reviewers (LQ, JAK) conducted an exercise of title and abstract review to independently include or exclude the first 200 citations. Inter-rater agreement for citation inclusion was calculated using percent agreement and 81% agreement was reached, meeting JBI recommendations (42) of 75% needed to proceed to the next stage of title and abstract review of all screened studies. No agreement calculations lower than reported herein were observed during the process. Discrepancies were resolved with an additional reviewer (AKQ). Where a decision could not be made between the three reviewers (LQ, JAK, AKQ) to include or exclude a citation, the three reviewers and two arbitrators (AR, CJG) reviewed the citation together to reach a resolution. Following completion of title and abstract review, full-text articles were retrieved for all included citations. The full text citations were subsequently assessed in detail against the inclusion criteria by two reviewers. Two reviewers (LQ, JAK) screened the first 100 full-text studies and reached 80% agreement.

### Data Items and Data Extraction Process

For full-text articles meeting inclusion criteria, reviewers extracted data independently and reviewed discrepancies. Due to the quantity of citations meeting inclusion criteria, additional coders were added to the research team and received training equivalent to the original coders. Data charting was completed in duplicates by seven independent reviewers (LQ, JAK, MC, MP, RM, SP, SR) and discrepancies were resolved with an additional reviewer (AKQ or RM). The extracted variables were conceptualized with the intent of capturing the breadth of sociodemographic identity domains (e.g. Race, Ethnicity, Gender, Sexual Orientation, Age, etc.) reported in the RCTs meeting inclusion criteria. After reviewing several articles and guidelines for sociodemographic reporting (28,32,33), the authors aligned on the inclusion of 16 demographic domains for data extraction (see Table 2). Citations that reported on secondary analysis of an RCT otherwise meeting inclusion criteria were excluded in this phase, and confirmation was subsequently provided that the primary citation was included for data extraction. A list of coded variables are provided in Supplemental 3. The final dataset of all included studies is available in Supplemental 4. Consistent with the guidance report developed by members of JBI and JBI Collaborating Centers, we did not conduct a critical appraisal of methodological quality or risk of bias of the included citations (46).

**Table 1.** Study Characteristics.

| **Study Characteristics (N = 299)** | **Count (%)** |
| --- | --- |
| *Year of Publication* |  |
| 2009 - 2013 | 5 (2%) |
| 2014 - 2018 | 65 (21%) |
| 2019 - 2022 | 229 (77%) |
| *Country of Origin* |  |
| Non-US | 194 (65%) |
| US | 105 (35%) |
| *Psychiatric Diagnoses* |  |
| Clinical Diagnosis | 177 (59%) |
| No Clinical Diagnosis | 122 (41%) |
| *Secondary Medical or Other Diagnoses* |  |
| Medical Diagnosis | 49 (16%) |
| No Medical Diagnosis | 250 (84%) |
| *Demographics Represented by Treatment Arm* | 256 (85%) |
| *Demographics Represented for Study Dropouts* | 20 (7%) |
| *Primary Outcomes Analyzed by Sociodemographic Category* | 36 (12%) |

**Table 2.** Number and Percentage of Studies that Reported Each Sociodemographic Domain.

| **Sociodemographic Domain (N = 299)** | **Count (%)** |
| --- | --- |
| Gender or Sex | 293 (98%) |
| Gender Only | 152 (51%) |
| Sex Only | 85 (28%) |
| Gender and Sex | 2 (<1%) |
| Gender/Sex Unspecified but Reported | 54 (18%) |
| Age | 290 (97%) |
| Education | 200 (67%) |
| Race or Ethnicity | 144 (48%) |
| Race Only | 41 (14%) |
| Ethnicity Only | 43 (14%) |
| Both Race and Ethnicity Distinctly Reported | 31 (10%) |
| Race or Ethnicity Unspecified but Reported | 29 (10%) |
| Employment | 127 (42%) |
| Marital Status | 116 (39%) |
| Income Level | 49 (16%) |
| Housing Situation | 19 (6%) |
| Residency or Location | 14 (5%) |
| Veteran Status | 11 (4%) |
| Number of Children | 8 (3%) |
| Sexual Orientation | 7 (2%) |
| Disability Status | 7 (2%) |
| Food Security | 1 (<1%) |

## RESULTS

### Study Characteristics

There were no included studies published in 2007 and 2008, thus the 299 studies included in the review were published between 2009 and 2022, with over three-quarters published after 2018 (Table 1). Approximately 35% were conducted in the US and 59% of the studies recruited a sample of participants with Diagnostic and Statistical Manual 5 (DSM-5) clinical diagnoses (47), with 16% including secondary medical or other physical diagnoses (e.g., chronic pain, multiple sclerosis, etc.) as part of inclusion criteria. The vast majority (85%) presented sociodemographics separately for each treatment arm, but few studies presented separate sociodemographics for study dropouts (7%), or analyzed the primary outcomes separately by different sociodemographic categories (12%).

### Primary Objectives

Frequency and Type of Sociodemographic Domains

On average across the 299 included studies, researchers reported 4.64 (SD = 1.79; range 0 - 9) of the 16 sociodemographic domains assessed in this review (see Table 2 and Figure 2 for the number and percentage of studies that reported data for each of the 16 sociodemographic domains included in this review). The most commonly reported sociodemographic domains reported by at least 50% of studies were Age (97%), Education (67%), and Gender (51%). The least common domains reported by 10% or fewer of studies were Housing Situation (e.g., living alone, with parents, homeless, etc; 6%), Residency/Location (e.g., rural vs urban; 5%), Veteran Status (4%), Number of Children (3%), Sexual Orientation (2%), Disability Status (2%), and Food Security (<1%).

**Figure 2.**
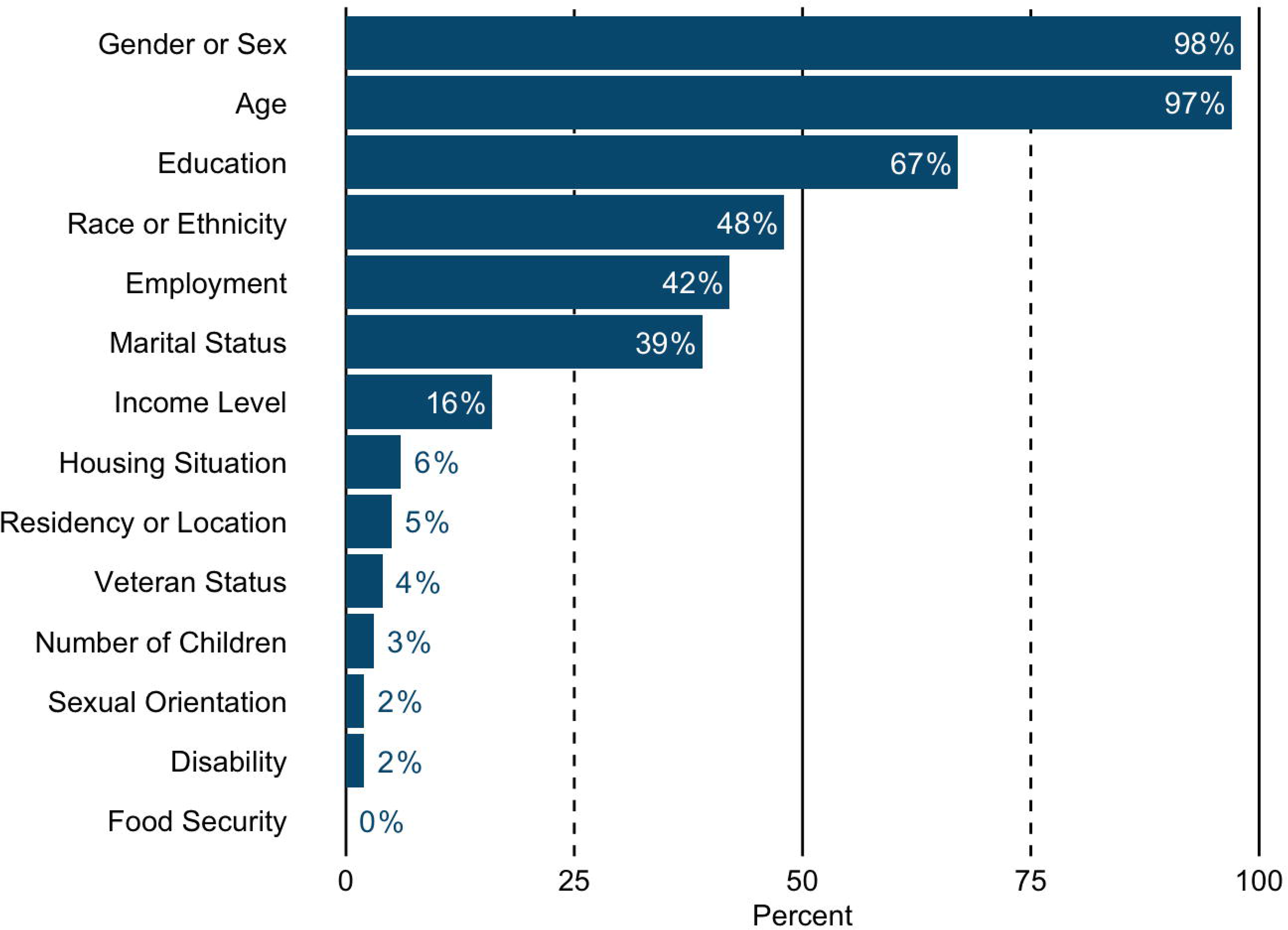
Percentage of studies that reported each sociodemographic domain. Note: Percentages were rounded to the nearest whole number; the domains of Gender or Sex, and Race or Ethnicity were combined in the figure for visual clarity.

Reporting of Gender/Sex and Race/Ethnicity Domains

Due to the observed common conflation of gender and sex, the domains of gender and sex were coded into the following buckets for clarity: “Gender Only”, “Sex Only”, “Gender and Sex”, and “Gender/Sex Unspecified but Reported” (see Figure 3). “Gender or Sex” represents the combination of these categories. “Gender and Sex” reflects instances in which the publication reported both sociodemographic domains separately. “Gender/Sex Unspecified but Reported” represents instances in which publications reported categories reflecting gender or sex, but did not specify which domain the category belonged to. For example, several of these studies included in their sociodemographic reporting the proportion of participants who were women, but did not indicate whether this was intended to capture participants’ gender or their sex. Following the same logic, the domains of Race and Ethnicity were coded into the following buckets: “Race Only”, “Ethnicity Only”, “Race and Ethnicity”, and “Race/Ethnicity Unspecified but Reported” (see Figure 4).

**Figure 3.**
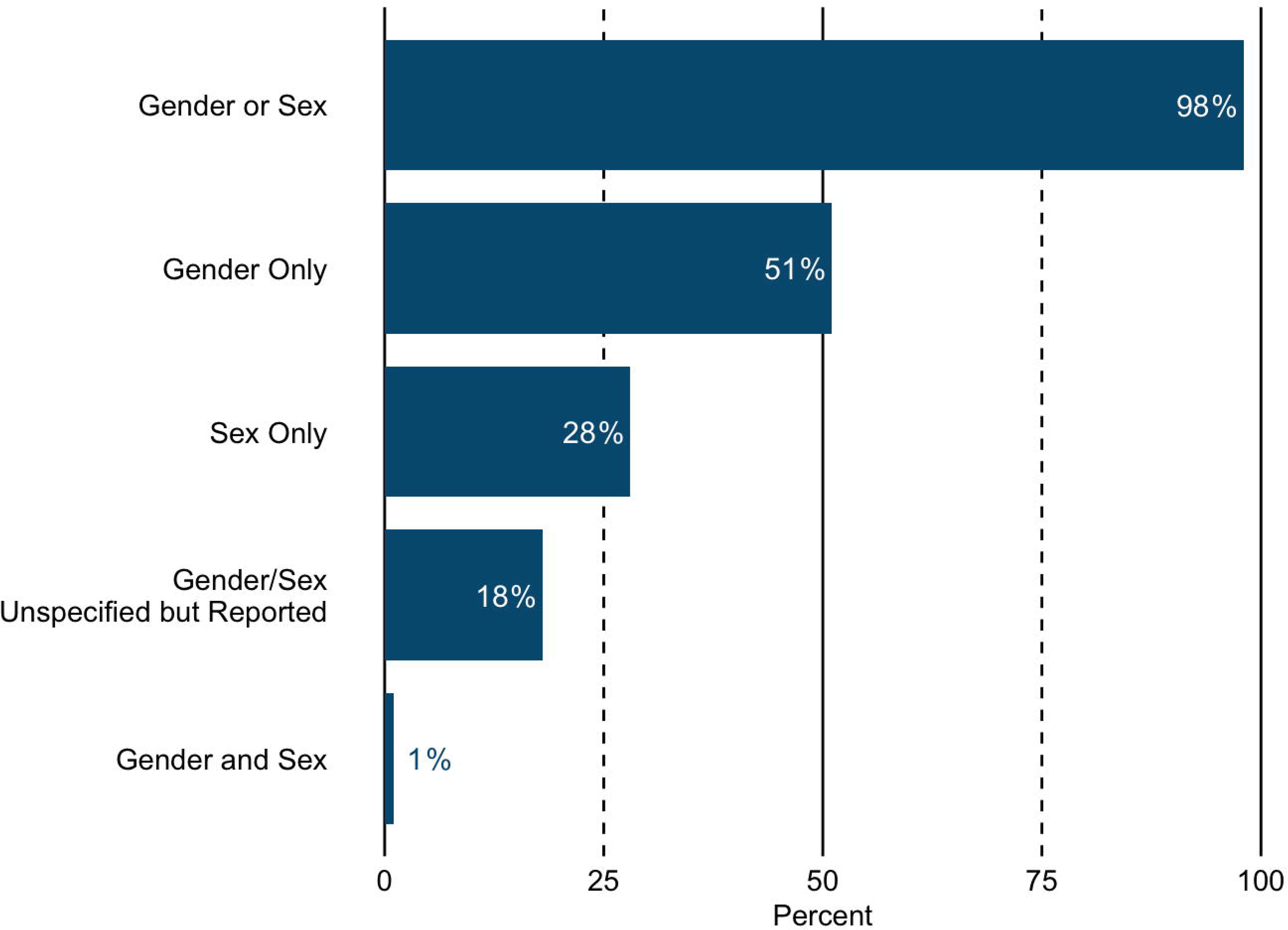
Percentage of studies that reported Gender and Sex domains.

**Figure 4.**
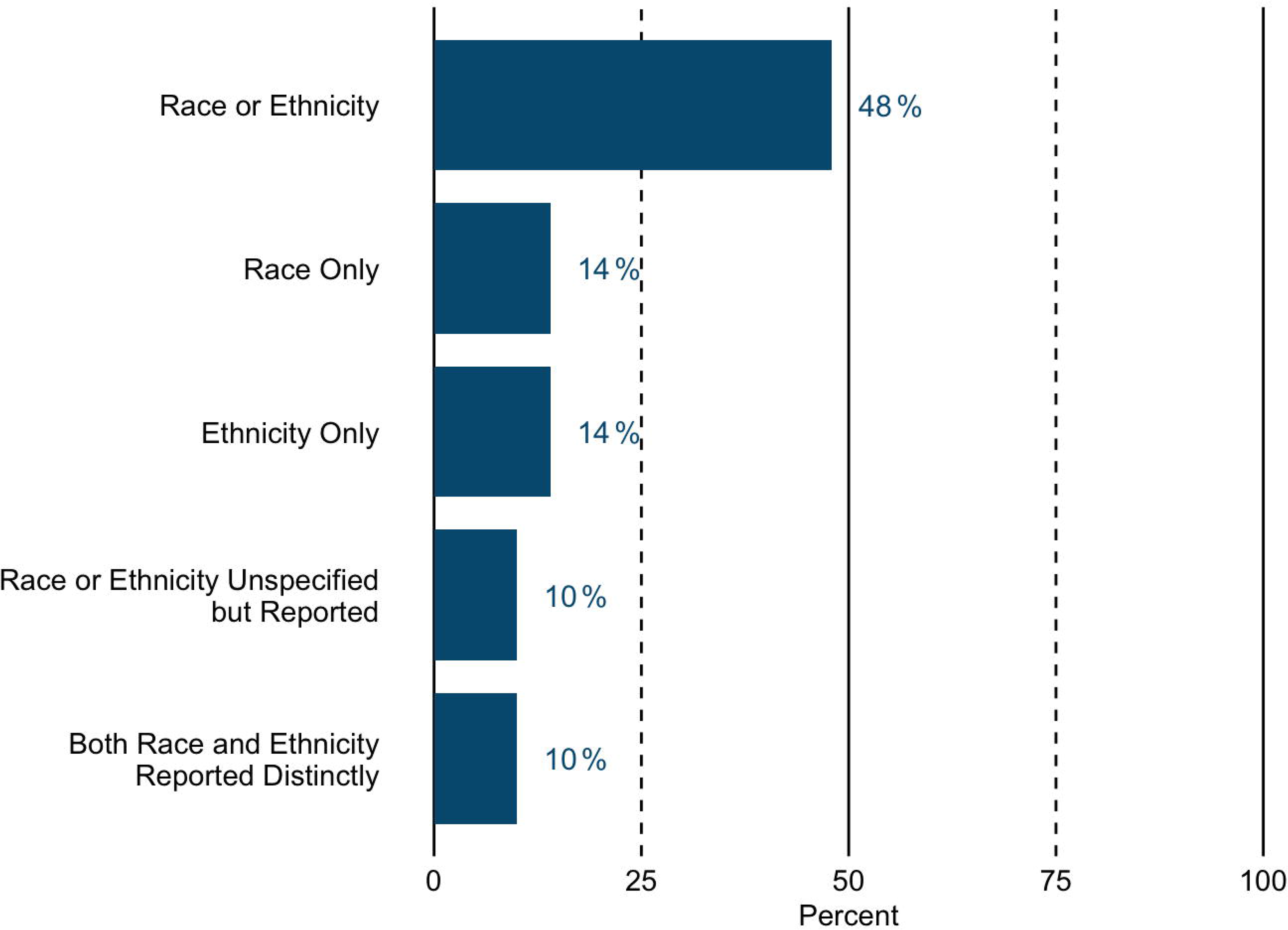
Percentage of studies that reported Race and Ethnicity domains.

Unlisted Sociodemographic Domains

To ensure the breadth of sociodemographic domains was thoroughly captured, the present review also documented domains that were not included in the 16 domains reviewed (called “Unlisted”). There were 35 studies (12%) that reported Unlisted sociodemographic domains. Five main clusters emerged: there were 10 studies that documented “Nationality”, 6 studies that documented “Parental Education”, 5 studies that documented “Languages”, 3 studies that documented “Religion”, and 3 studies that documented “Subjective Socioeconomic Status”.

Categories Reported within Sociodemographic Domains

Table 3 provides descriptives for the categories reported within each sociodemographic domain (i.e., education, income, etc), including the most frequently reported number of categories (Mode), the highest and lowest reported number of categories (Range) and the two most common category options reported.

**Table 3.**
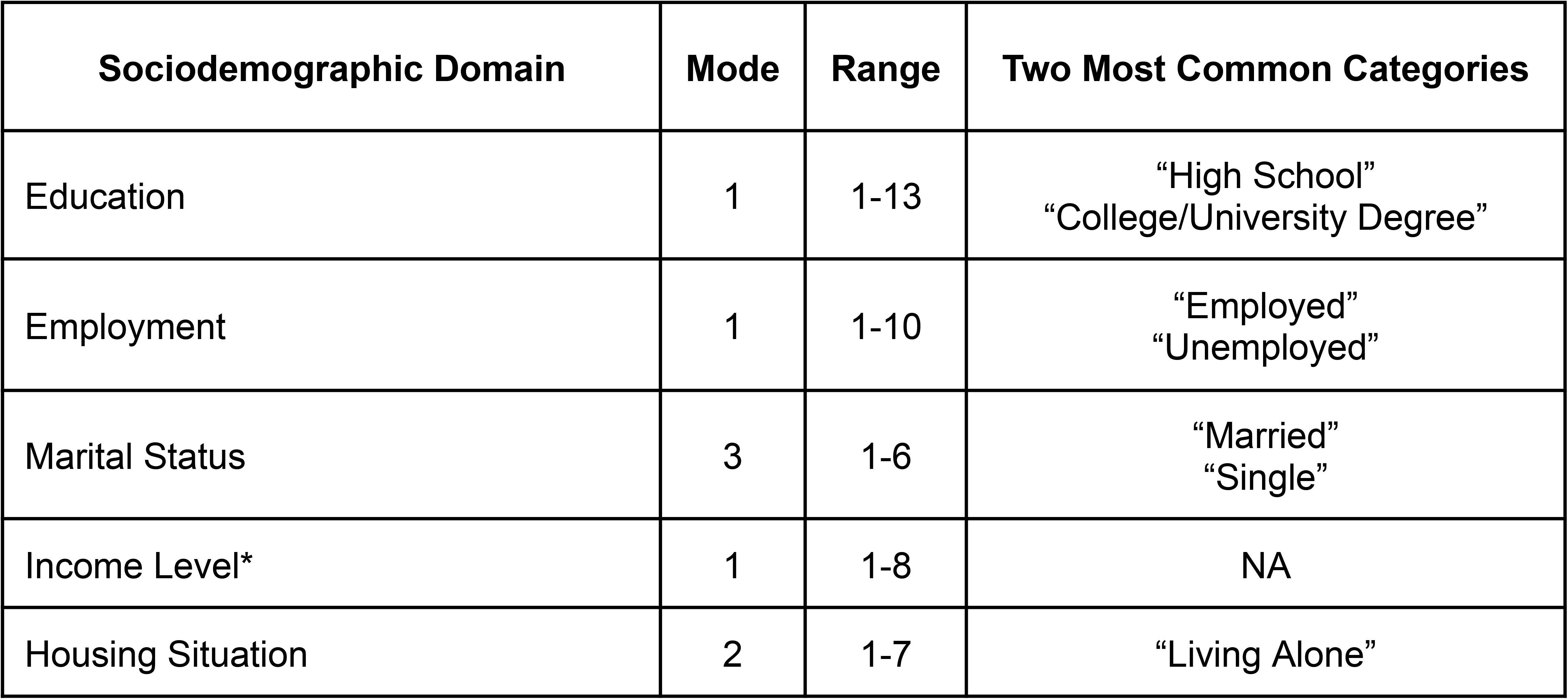
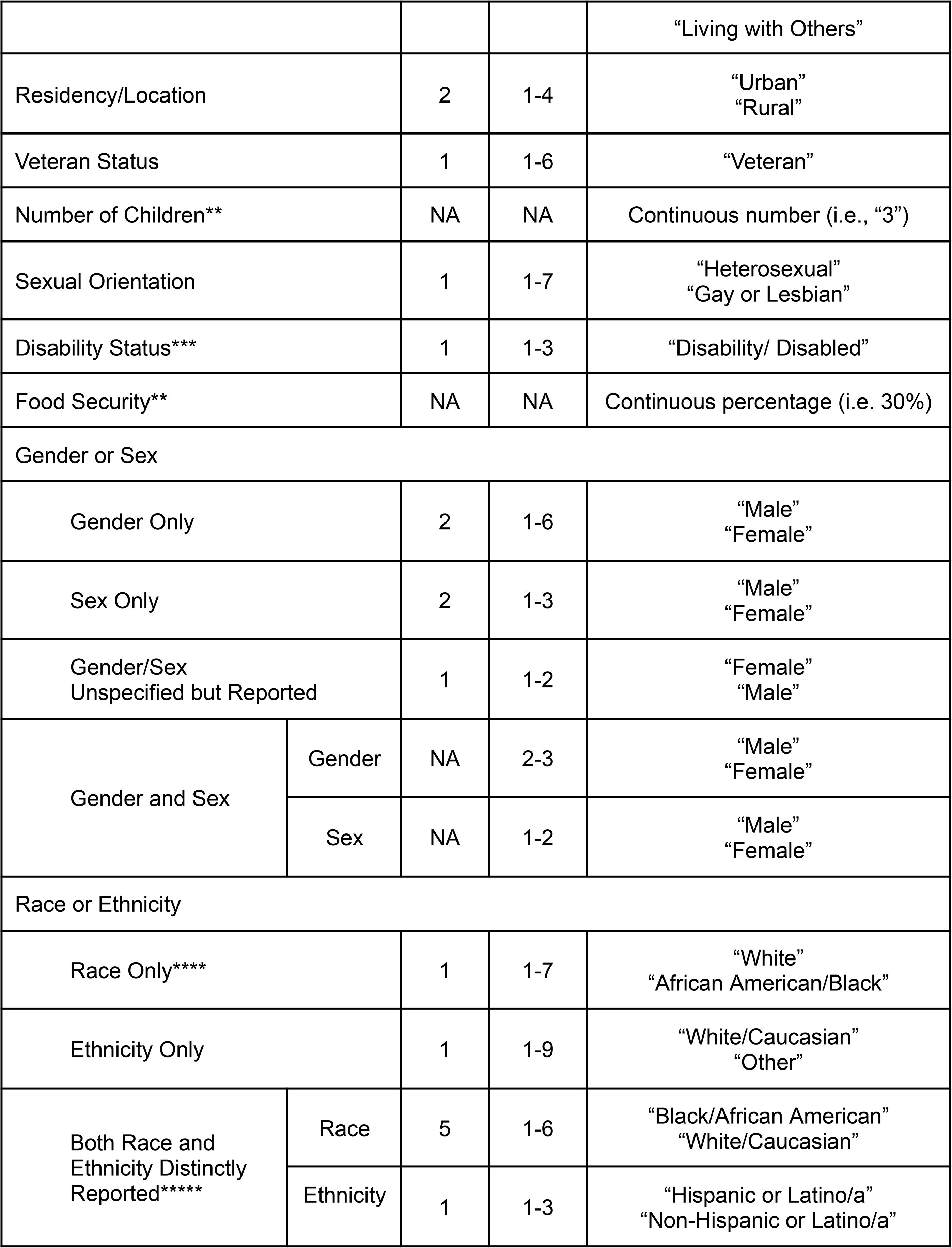
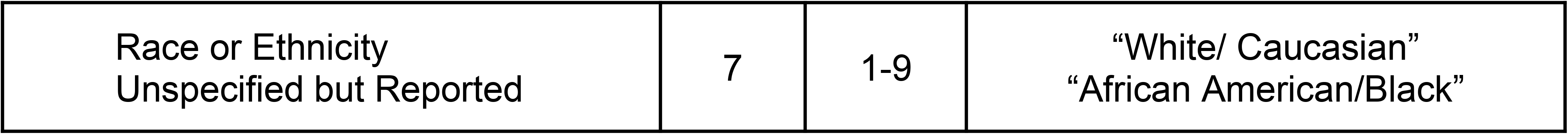
Number of Categories Reported Per Sociodemographic Domain.

| **Sociodemographic Domain** | **Mode** | **Range** | **Two Most Common Categories** |
| --- | --- | --- | --- |
| Education | 1 | 1-13 | “High School”<br>“College/University Degree” |
| Employment | 1 | 1-10 | “Employed”<br>“Unemployed” |
| Marital Status | 3 | 1-6 | “Married”<br>“Single” |
| Income Level* | 1 | 1-8 | NA |
| Housing Situation | 2 | 1-7 | “Living Alone” |

|  |  |  |  |  |
| --- | --- | --- | --- | --- |
|  |  |  |  | "Living with Others" |
| Residency/Location |  | 2 | 1-4 | "Urban"<br>"Rural" |
| Veteran Status |  | 1 | 1-6 | "Veteran" |
| Number of Children** |  | NA | NA | Continuous number (i.e., "3") |
| Sexual Orientation |  | 1 | 1-7 | "Heterosexual"<br>"Gay or Lesbian" |
| Disability Status*** |  | 1 | 1-3 | "Disability/ Disabled" |
| Food Security** |  | NA | NA | Continuous percentage (i.e. 30%) |
| Gender or Sex |  |  |  |  |
| Gender Only |  | 2 | 1-6 | "Male"<br>"Female" |
| Sex Only |  | 2 | 1-3 | "Male"<br>"Female" |
| Gender/Sex<br>Unspecified but Reported |  | 1 | 1-2 | "Female"<br>"Male" |
| Gender and Sex | Gender | NA | 2-3 | "Male"<br>"Female" |
|  | Sex | NA | 1-2 | "Male"<br>"Female" |
| Race or Ethnicity |  |  |  |  |
| Race Only**** |  | 1 | 1-7 | "White"<br>"African American/Black" |
| Ethnicity Only |  | 1 | 1-9 | "White/Caucasian"<br>"Other" |
| Both Race and<br>Ethnicity Distinctly<br>Reported***** | Race | 5 | 1-6 | "Black/African American"<br>"White/Caucasian" |
|  | Ethnicity | 1 | 1-3 | "Hispanic or Latino/a"<br>"Non-Hispanic or Latino/a" |
| Race or Ethnicity<br>Unspecified but Reported | 7 | 1-9 | "White/ Caucasian"<br>"African American/Black" |

Note.
\* For "Income Level", nearly all category options included a numeric range of salary (e.g., "Less than \$25,000", "\$25,001 - \$49,999", etc.) however the particular cutoffs varied widely and were not easily interpretable for summarization here.
\*\* The mode and range were not calculable ("NA") for "Number of Children" and "Food Security", as these domains were reported with a single continuous number or percentage.
\*\*\* For "Disability Status", the most frequently reported option was a variation of "disability" or "disabled", though we are combining across several variations for simplicity of reporting due to the small sample size.
\*\*\*\* For "Race Only" and "Race or Ethnicity Unspecified but Reported", for category "African American/Black", we combined across categories including "African American", "African", "Black", and "Black or African American".

### Secondary Objectives

Reporting of Sociodemographic Domains Over Time

To determine whether there were changes in reporting of sociodemographic domains over time, studies were divided into three sub-groups, each covering 4-5 years from 2009 to 2022, and frequency of domain reported was described (Table 4). There was an increase in total publications across time, with 5 studies published between 2009 and 2013, 65 between 2014 and 2018, and 229 from between 2019 and 2022. Overall, the descriptive trend suggests an increase in publications reporting sociodemographic domains in more recent years and also a decrease over time in both the number of studies which reported on Ethnicity Only (i.e., not Race), and those that reported but did not specify Gender or Sex.

**Table 4.** Number and Percentage of All Studies Reporting Sociodemographic Domains by 4-5 Year Time Period.

| Demographic Domain | 2009 - 2013<br>(n = 5) | 2014 - 2018<br>(n = 65) | 2019 - 2022<br>(n = 229) |
| --- | --- | --- | --- |
| Age | 4 (80%) | 63 (97%) | 223 (97%) |
| Gender or Sex | 4 (80%) | 65 (100%) | 224 (98%) |
| Gender Only | 0 | 33 (51%) | 119 (52%) |
| Sex Only | 0 | 20 (31%) | 65 (28%) |
| Gender and Sex | 0 | 0 | 2 (1%) |
| Gender/Sex Unspecified | 4 (80%) | 12 (18%) | 38 (17%) |
| Education | 1 (20%) | 47 (72%) | 152 (66%) |
| Race or Ethnicity | 2 (40%) | 36 (55%) | 106 (46%) |
| Race Only | 0 | 8 (12%) | 33 (14%) |
| Ethnicity Only | 2 (40%) | 13 (20%) | 28 (12%) |
| Both Race and Ethnicity Distinctly Reported | 0 | 6 (9%) | 25 (11%) |
| Race or Ethnicity Unspecified | 0 | 9 (14%) | 20 (9%) |
| Employment | 2 (40%) | 24 (37%) | 101 (44%) |
| Marital Status | 0 | 27 (42%) | 89 (39%) |
| Income Level | 0 | 10 (15%) | 39 (17%) |
| Residency of Location | 2 (40%) | 2 (3%) | 10 (4%) |
| Veteran Status | 0 | 6 (9%) | 5 (2%) |
| Number of Children | 0 | 1 (2%) | 7 (3%) |
| Housing Situation | 0 | 4 (6%) | 15 (7%) |
| Sexual Orientation | 0 | 0 | 7 (3%) |
| Disability Status | 0 | 1 (2%) | 6 (3%) |
| Food Security | 0 | 0 | 0 |

Sociodemographic Reporting: US vs. Non-US Studies

Reporting of sociodemographic domains was fairly consistent across US and non-US based studies (see Table 5). However, several domains appeared to differ, including Race, Ethnicity, Employment, and Veteran Status. Ninety percent of US studies reported on Race or Ethnicity, whereas 25% of non-US studies did so. US studies appear to have reported more frequently than non-US studies for Race Only (31% vs. 4%), Both Race and Ethnicity (27% vs. 2%), and Race or Ethnicity Unspecified (24% vs. 2%). However, US studies appear to have reported less frequently for Ethnicity Only (9% vs. 18%). For Employment, 46% of non-US studies reported, compared to 36% of US studies. Finally, Veteran Status was reported by 10% of US studies, but less than 1% of non-US studies.

**Table 5.** Comparison of Sociodemographic Domain Reporting Across US and Non-US Studies.

| **Demographic Domain** | **US Count (%)**<br>**(n = 105)** | **Non-US Count (%)**<br>**(n = 194)** |
| --- | --- | --- |
| Gender or Sex | 104 (99%) | 189 (97%) |
| Gender Only | 50 (48%) | 102 (53%) |
| Sex Only | 27 (26%) | 58 (30%) |
| Gender and Sex | 2 (2%) | 0 (0%) |
| Gender/Sex Unspecified but Reported | 25 (24%) | 29 (15%) |
| Age | 100 (95%) | 190 (98%) |
| Education | 71 (68%) | 129 (66%) |
| Race or Ethnicity | 95 (90%) | 49 (25%) |
| Race Only | 33 (31%) | 8 (4%) |
| Ethnicity Only | 9 (9%) | 34 (18%) |
| Both Race and Ethnicity Distinctly Reported | 28 (27%) | 3 (2%) |
| Race or Ethnicity Unspecified but Reported | 25 (24%) | 4 (2%) |
| Employment | 38 (36%) | 89 (46%) |
| Marital Status | 42 (40%) | 74 (38%) |
| Income Level | 26 (25%) | 23 (12%) |
| Residency/Location | 5 (5%) | 9 (5%) |
| Veteran Status | 10 (10%) | 1 (1%) |
| Number of Children | 1 (1%) | 7 (4%) |
| Housing Situation | 6 (6%) | 13 (7%) |
| Sexual Orientation | 4 (4%) | 3 (2%) |
| Disability Status | 3 (3%) | 4 (2%) |
| Food Security | 1 (1%) | 0 (0%) |

## DISCUSSION

Our comprehensive scoping review of 299 RCTs of app-based DMHIs published from 2007 to 2022 found that this research underreported key sociodemographic data. Across the 16 sociodemographic domains reviewed, the average number of domains reported per study was 4.6, and no domains were consistently reported across studies. Three domains (Age, Education, Gender) were reported in greater than 50% of studies and seven domains (Housing Situation, Residency/Location, Veteran Status, Number of Children, Sexual Orientation, Disability, and Food Security) were reported by fewer than 10% of studies. The remaining domains (Sex, Race, Ethnicity, Employment, Marital Status, and Income Level) were reported between 10% and 42% of the time.

Reporting practices for the domains of Gender, Sex, and Sexual Orientation demonstrated widespread shortcomings, including limited reporting and apparent conflation across these domains. Gender and Sex were commonly used interchangeably; however, these terms represent distinct definitions in contemporary science (48). Sex refers to the biologically assigned classification of male or female according to genetic, anatomical, and hormonal characteristics (48,49). Gender identity refers to socially constructed roles, behaviors, and identities generally (but not necessarily) associated with an individual’s Sex (48,49). Importantly, Gender may not align with an individual’s assigned sex at birth and individuals may identify as transgender, genderqueer, non-binary, and other genders that do not conform to traditional categories. Recognition of this is evident in only two studies that reported both Sex and Gender as separate, distinct domains. Moreover, the most frequently reported number of category options for Gender and Sex was two, and these were “Male” and “Female” for both, suggesting that researchers are regularly conflating Gender and Sex. Additionally, fewer than 1 in 20 studies reported options inclusive of the broader spectrum of gender identities, such as transgender man/woman, genderfluid, or non-binary, indicating that the vast majority of researchers are omitting inclusive options. Finally, there was a near total absence of reporting on Sexual Orientation, with just 2% of studies reporting on this domain. The omission of reporting for sexual and gender minorities has also been demonstrated by Kimber and colleagues (25) in a review of psychological interventions for physical health, and by Pachinkis and colleagues (2018) in a review of lesbian, gay, bisexual, transgender, and queer (LGBTQ+) affirming mental health care (50). Despite these gaps in reporting and representation, it has become clear that the LGBTQ+ community experiences social stigma, rejection, violence, and discrimination; all factors that contribute to high rates of depression, anxiety, substance use, and suicidal ideation (4,51–53). Fortunately, these pressing issues are gaining wider recognition; for example, in 2021 the US Census Bureau attempted for the first time to collect data on LGBTQ+ Americans in a large real-time national survey (54). These efforts, while modest, are encouraging. The continued collection and reporting on Sexual Orientation and Gender Identity is essential to serving members of the LGBTQ+ community.

Race and Ethnicity were also not widely reported, regularly conflated, and limited in their options for self-identification. Fewer than half of studies reported on either Race or Ethnicity at all, and of those that did, reporting practices were varied and inconsistent. Following the most recent 2021 guidance published by Flanagin and colleagues (29), race and ethnicity are defined as social constructs that are not biologically based and are used to characterize human populations based on shared physical characteristics, cultural, geographical factors. Specifically, race typically refers to a social construct that categorizes people based on physical characteristics, whereas ethnicity refers to a shared cultural heritage, geographical origin, and ancestry. The lack of specificity observed in many studies appears to have resulted in a profusion of category options mixed indiscriminately across both domains, such as “White/Caucasian” (which typically refers to race) appearing under Ethnicity, and “Hispanic” (which typically refers to ethnicity) appearing as an option under Race. Nearly all of the studies that did distinctly report both Race and Ethnicity were published in the United States after 2018, which may reflect the influence of the recent US Food and Drug Administration (FDA) and US Census Bureau guidelines (55,56). Relatedly, we observed that reporting practices regarding Race and Ethnicity may have differed between the US and non-US countries. Studies published in the US were fairly consistent in reporting at least one of Race or Ethnicity (90%); one-quarter of non-US studies reported the same. This may be due in part to international variations in racial and ethnic diversity, which may correspond to differing reporting practices and guidelines. For example, guidelines published by the Institute for Clinical and Economic Review (ICER), recognize the possibility of barriers to recruitment of diverse racial and ethnic samples in multinational trials, and as such only apply racial and ethnic diversity ratings to US-based trials (57).

Nonetheless, continued improvement in data collection and reporting practices regarding race and ethnicity is critical, especially in the US, where members of marginalized racial and ethnic groups show higher rates of being undiagnosed or underdiagnosed for mental health conditions (58) and barriers such as stigma, discrimination, and lack of access to care further contribute to hesitancy in seeking mental healthcare (59–63).

On a positive note, trends suggested that the frequency of reporting across sociodemographic domains has generally increased over time, especially in recent years from 2018 onward. A number of converging factors may play a role in this trend, including efforts put forth by advocacy and special interest groups, advances in social justice movements, growing awareness and emphasis of diversity in scientific conferences and educational curricula, as well as publication of guidelines and funding requirements by organizations such as the NIH and FDA. The continuation of these trends toward improvement in sociodemographic reporting will be critical to understand the impact of DMHI across diverse populations.

### Strengths and Limitations

This study had several strengths. It is the first of its kind to assess sociodemographic reporting practices in RCTs of app-based DMHIs, and lays the foundation for improvement of sociodemographic data collection and reporting in future studies. This review was both large and comprehensive, screening over 5000 article abstracts leading to the inclusion of nearly 300 articles spanning the entire lifespan of app-based DMHIs and extracting data for a wide array of 16 sociodemographic domains. Article inclusion criteria allowed for review of a broad range of DMHI studies, including global populations with both clinical and sub-clinical conditions. There were also several limitations. This scoping review did not describe the actual composition of each study sample due to the large number of studies included in the review and the breadth of domains evaluated within. This review was also merely descriptive; quantitative analyses such as formal assessment of statistical differences across reporting practices or time periods were beyond the review’s scope, and results should be interpreted accordingly.

## Conclusion

This scoping review describes widespread underreporting of sociodemographic information in RCTs of app-based DMHIs published globally from 2007 to 2022. No sociodemographic domain was consistently reported across all 299 studies and reporting was often incomplete (i.e., % female only), unclear (i.e., conflation of Gender and Sex; Race and Ethnicity), and limited (i.e., only options representing majority groups were offered). Trends did suggest improvements in reporting in recent years, which may be a result of ongoing and growing efforts and awareness brought to issues of diversity, equity, and inclusion in science and healthcare.

All research, including DMHI, optimally invites and studies the personal, social, cultural, environmental, and economic variables that shape the lives of study participants to determine how such sociodemographic domains interact with, predict, and facilitate mental health outcomes and other key variables of interest (i.e., treatment satisfaction). Thorough collection and reporting of these sociodemographic data will necessarily create opportunities for enhanced learning and understanding of diverse populations. Equipped with such knowledge and data, DMHI researchers may outline the potential generalizability of their results as well as create and study treatment adaptations as indicated. Ultimately, improved sociodemographic collection and reporting practices would enhance the diversity of DMHI literature overall and also better position DMHIs to facilitate reducing disparities in mental healthcare.

## CONTRIBUTIONS

AKQ in collaboration with AR, PW, AD, JAK, LQ, and CJG, conceptualized and designed the study and drafted the protocol. JAK and LQ, in coordination with AKQ, conducted the database search, title/abstract screening, and full-text screening. AKQ, JAK, and LQ, with support from MC, MP, RM, SP, and SR completed data extraction. AR and CJG moderated discrepancy resolution throughout. Data synthesis and analysis were completed by AKQ, RM, and MC. MC, MP, AR, and CJG drafted the background section. JAK drafted the methods section and completed Supplemental 2 (search strings/terms to identify included papers) and 3 (variable descriptions). JAK, RM, and AKQ created Figure 1 and PRISMA-ScR requirements (Supplemental 1). RM and MC drafted the results and discussion sections. MP prepared the reference section and RM prepared the final dataset (Supplemental 4). All authors contributed to multiple draft revisions, with ownership for incorporation of all author edits and flow of overall manuscript championed by RM, MC, and AR. All authors read and approved the submitted version.

We would like to thank Adam Platt, MSc, for coordinating scientific communications and facilitating the organization of the manuscript preparation and submission.

## DATA SHARING STATEMENT

The dataset underlying the results of this study is published in Supplemental 4.

## Supporting information

Supplemental 1 PRISMA Checklist

Supplemental 2 Search Strings

Supplemental 3 Variable Descriptions

## Data Availability

All data produced in the present study will be available as a supplement to the published article.

