## Supplemental 2 Search Strings for "Common Practices for Sociodemographic Data Reporting in Digital Mental Health Intervention Research: A Scoping Review"

### Supplemental 2 (PRISMA #8) Search strings

**Appendix 2: Search Terms to Identify Included Papers**

**Final Search Strings**

Database: PubMED <English language articles only, January 1 2007 to August 9, 2022>; EBSCO hosted CINAHL and PsycINFO <choose databases ‘CINAHL Plus with Full Text’ and ‘APA PsycINFO’, uncheck ‘apply equivalent subjects,’ English language articles only, ‘Published Date’ January 1 2007 to August 9, 2022; PsycInfo special limiters, 2007 start year for ‘Publication Year,’ English ‘Language’; CINAHL special limiters, English ‘Language’>; Google Scholar < ‘Return articles dated between’ 2007 lower boundary>; PsyArXiv <database without search filters>. Search conducted August 9, 2022.

**PubMED**

("mental disorders"[MeSH Terms] OR "anxi*"[tiab] OR "agoraphobia"[tiab] OR "panic"[tiab] OR "phobia"[tiab] OR "post-traumatic stress"[tiab] OR "mental illness*"[tiab] OR "mental health"[tiab] OR "affective disorder*"[tiab] OR "psychiatric disorder*"[tiab] OR "bipolar"[tiab] OR "mania"[tiab] OR "mood disorder*"[tiab] OR "depress*"[tiab] OR "seasonal affective disorder"[tiab] OR "psychosis"[tiab] OR "psychotic"[tiab] OR "schizophr*"[tiab] OR "well-being"[tiab] OR "wellbeing"[tiab] OR "quality of life"[tiab] OR "self-harm*"[tiab] OR "self-injury"[tiab] OR "stress*"[tiab] OR "distress*"[tiab] OR "mood"[tiab] OR "body image" [tiab] OR "eating disorder*"[tiab] OR “binge-eating”[tiab] OR “insomnia”[tiab] OR “bulimi*”[tiab] OR "sleep disorder*"[tiab] OR "sleep disturbance"[tiab] OR “anorexia”[tiab] OR "sleep problem*"[tiab] OR "traumatic"[tiab] OR "suicid*"[tiab] OR "addic*"[tiab] OR “substance use”[tiab] OR “alcohol abuse”[tiab] OR “alcohol depend*”[tiab] OR “psychological”[tiab] OR “psychology”[tiab] OR “behavioral”[tiab] OR “mindfulness”[tiab] OR “oppositional defiant disorder”[tiab] OR “fatigue”[tiab] OR “self-compassion”[tiab]) AND ("mobile applications"[MeSH Terms] OR "smartphone"[ti] OR "mobile phone"[ti] OR "cell phone"[ti] OR "app"[ti] OR "iphone"[ti] OR "android"[ti] OR "mhealth"[ti] OR "m-health"[ti] OR "mobile health"[ti] OR "mobile app"[ti] OR "phone application"[ti] OR "mobile device*"[ti] OR "mobile-based"[ti] OR "cellular phone"[ti]) AND ("Clinical Trial"[Publication Type] OR "random*"[ti] OR "trial*"[ti] OR "allocat*"[ti] OR "random assignment"[ti] OR "controlled"[ti] OR "clinical trial"[ti] OR "control group"[ti])

**CINAHL & PsycINFO**

("mental disorders" OR "anxi*" OR "agoraphobia" OR "panic" OR "phobia" OR "post-traumatic stress" OR "mental illness*" OR "mental health" OR "affective disorder*" OR "psychiatric disorder*" OR "bipolar" OR "mania" OR "mood disorder*" OR "depress*" OR "seasonal affective disorder" OR "psychosis" OR "psychotic" OR "schizophr*" OR "well-being" OR "wellbeing" OR "quality of life" OR "self-harm*" OR "self-injury" OR "stress*" OR "distress*" OR "mood" OR "body image" OR "eating disorder*" OR “binge-eating” OR “insomnia” OR “bulimi*” OR "sleep disorder*" OR "sleep disturbance" OR “anorexia” OR "sleep problem*" OR "traumatic" OR "suicid*" OR "addic*" OR “substance use” OR “alcohol abuse” OR “alcohol depend*” OR “psychological” OR “psychology” OR “behavioral” OR “mindfulness” OR “oppositional defiant disorder” OR “fatigue” OR “self-compassion”) AND ("mobile applications" OR "smartphone" OR "mobile phone" OR "cell phone" OR "app" OR "iphone" OR "android" OR "mhealth" OR "m-health" OR "mobile health" OR "mobile app" OR "phone application" OR "mobile device*" OR "mobile-based" OR "cellular phone") AND ("Clinical Trial" OR "random*" OR "trial*" OR "allocat*" OR "random assignment" OR "controlled" OR "clinical trial" OR "control group")

**Google Scholar**

("mental disorders" OR anxi* OR "mental illness*" OR "mental health" OR "mood disorder*" OR depress* ) AND ("mobile applications" OR smartphone OR "mobile phone" OR mhealth OR "mobile health" OR app) AND ("clinical trial" OR "randomized controlled trial")

**PsyArXiv**

("mental disorders" OR "anxi*" OR "agoraphobia" OR "panic" OR "phobia" OR "post-traumatic stress" OR "mental illness*" OR "mental health" OR "affective disorder*" OR "psychiatric disorder*" OR "bipolar" OR "mania" OR "mood disorder*" OR "depress*" OR "seasonal affective disorder" OR "psychosis" OR "psychotic" OR "schizophr*" OR "well-being" OR "wellbeing" OR "quality of life" OR "self-harm*" OR "self-injury" OR "stress*" OR "distress*" OR "mood" OR "body image" OR "eating disorder*" OR “binge-eating” OR “insomnia” OR “bulimi*” OR "sleep disorder*" OR "sleep disturbance" OR “anorexia” OR "sleep problem*" OR "traumatic" OR "suicid*" OR "addic*" OR “substance use” OR “alcohol abuse” OR “alcohol depend*” OR “psychological” OR “psychology” OR “behavioral” OR “mindfulness” OR “oppositional defiant disorder” OR “fatigue” OR “self-compassion”) AND ("mobile applications" OR "smartphone" OR "mobile phone" OR "cell phone" OR "app" OR "iphone" OR "android" OR "mhealth" OR "m-health" OR "mobile health" OR "mobile app" OR "phone application" OR "mobile device*" OR "mobile-based" OR "cellular phone") AND ("Clinical Trial" OR "random*" OR "trial*" OR "allocat*" OR "random assignment" OR "controlled" OR "clinical trial" OR "control group")
