## Supplemental 3 Variable Descriptions for "Common Practices for Sociodemographic Data Reporting in Digital Mental Health Intervention Research: A Scoping Review"

### Supplemental 3 (PRISMA #11) Variable descriptions

**Appendix 3: All variables for which data were sought and simplifications made.**

**Scoping review title:** Common Practices for Sociodemographic Data Reporting in Digital Mental Health Intervention Research: A Scoping Review

**Objective:** Determine common practices for collection of sociodemographic information in randomized clinical trials involving app-based psychological interventions.

**Inclusion criteria:** Randomized control trials, mobile app-based intervention focused on mental health, journal articles published as of January 1, 2007 or after, English language articles.

**Population:** Any

**Concept:** Collection of sociodemographic domain variables in digital mental health-focused randomized control trials.

**Context:** Randomized control trials

**Exclusion criteria:** Not a randomized control trial, not a journal article, intervention is not primarily mobile-app based, intervention is not mental health focused. During the full-text phase non-journal articles were excluded including protocols, conference abstracts, clinical trial registrations, dissertations, and erratums that did not relate to sociodemographic domains. Secondary analyses were excluded in the data extraction phase.

**Data Extraction Categories**

1. Author list
2. Publication Year
3. Journal
4. First author last name-Year
5. Title
6. Primary study?
7. Exclude at Data Extraction?
8. Notes
9. Intervention
10. Cont1
11. Cont2
12. Population
13. Target psych disorder
14. Non-clinical specifier
15. Medical/other Diagnosis
16. Country of origin
17. Age reported?
18. Gender reported?
19. Gender categories
20. Sex reported?
21. Sex categories
22. Gender/sex unspecified but reported?
23. Gender/sex unspecified categories
24. Sexual orientation reported?
25. Sexual orientation categories
26. Race reported?
27. Race categories
28. Ethnicity reported?
29. Ethnicity categories
30. Marital status reported?
31. Marital status categories
32. Education reported?
33. Education categories
34. Employment reported?
35. Employment categories
36. Income level reported?
37. Income level categories
38. Disability reported?
39. Disability categories
40. Veteran status reported?
41. Veteran status categories
42. Number of children reported?
43. Housing situation reported?
44. Housing situation categories
45. Food security reported? y/n
46. Residency/location reported?
47. Residency/location categories
48. Unlisted Category reported?
49. Unlisted Category
50. Demographics reported by treatment arm?
51. Demographics reported for study dropouts?
52. Primary outcomes broken down by sociodemographic category?
